# Transfer learning applied to the forecast of mosquito-borne diseases

**DOI:** 10.1101/2020.02.03.20020164

**Authors:** Flávio Codeço Coelho, Nicolaus Linneu de Holanda, Beatriz Coimbra

**Affiliations:** School of Applied Mathematics, Getulio Vargas Foundation, Rio de Janeiro, RJ, Brazil; Department of Condensed Matter, Applied Physics and Nanoscience, Brazilian Center for Research in Physics, Rio de Janeiro, RJ, Brazil

## Abstract

Here we apply the concept of transfer learning to time series forecasting models for mosquito-borne diseases. Transfer learning, in this application, allows us to use knowledge obtained from modeling one disease to predict an emerging one for which extensive data is still not available. Here we discuss the performances of two families of models for predicting Chikungunya and Zika using models trained with dengue time series, in two Brazilian cities: Rio de Janeiro and Fortaleza.

## Introduction

It is common for a given insect to be the main vector for more than one pathogen. this is certainly true for the mosquito Aedes aegypti, which is a vector for various viruses such as the dengue virus (DENV), Zika Virus (ZIKV), Chikungunya virus (CHIKV), yellow fever virus (YFV) among others. It is also common to have a lot of historical transmission data on one disease but little to none on a emerging one.

Aedes aegypti’s distribution is restricted to tropical and subtropical climates, which reflects its sensitivity to temperature, humidity and other weather constraints. The modulation of its life cicle by climate, shapes the seasonality of the diseases it transmits. In Brazil, Aedes aegypti has been historically associated with the transmission of dengue, making it a marked seasonal disease. In recent years, A. aegypti has also been notably responsible for epidemics of the Zika and Chikungunya virus.

In this paper we propose to adopt the widely used method of transfer-learning to take advantage of the long time series available for dengue and explore the performance of dengue forecast models to predict the weekly incidences of Zika and Chikungunya as well. Transfer learning is a method used in association with deep learning models. Here we will use a deep learning model, in the form of a LSTM (long short term memory) recursive neural network model, which we have shown, in a previous work[ref], to yield accurate forecasts for weekly dengue incidence. But we will also use an ensemble regression model, namely a Random Quantile Forest model. Besides weekly incidence, climate variables such as temperature, humidity, and atmospheric pressure are also used as predictors. A spatial component built from the incidence at neighboring cities is also included. We present results of the forecast of total incidence of arboviral disease as well as of each disease separately and discuss the relative performances of the model for each of these tasks.

## Materials and methods

### Data

The data used in this work comes from the Infodengue project [1], which monitors these arboviroses in Brazil. In figure 1 we can see the incidence series for states in Brazil which had significant outbreaks of the 3 arboviroses since 2016 soon after Chikungunya and Zika arrived in Brazil.

**Fig 1.**
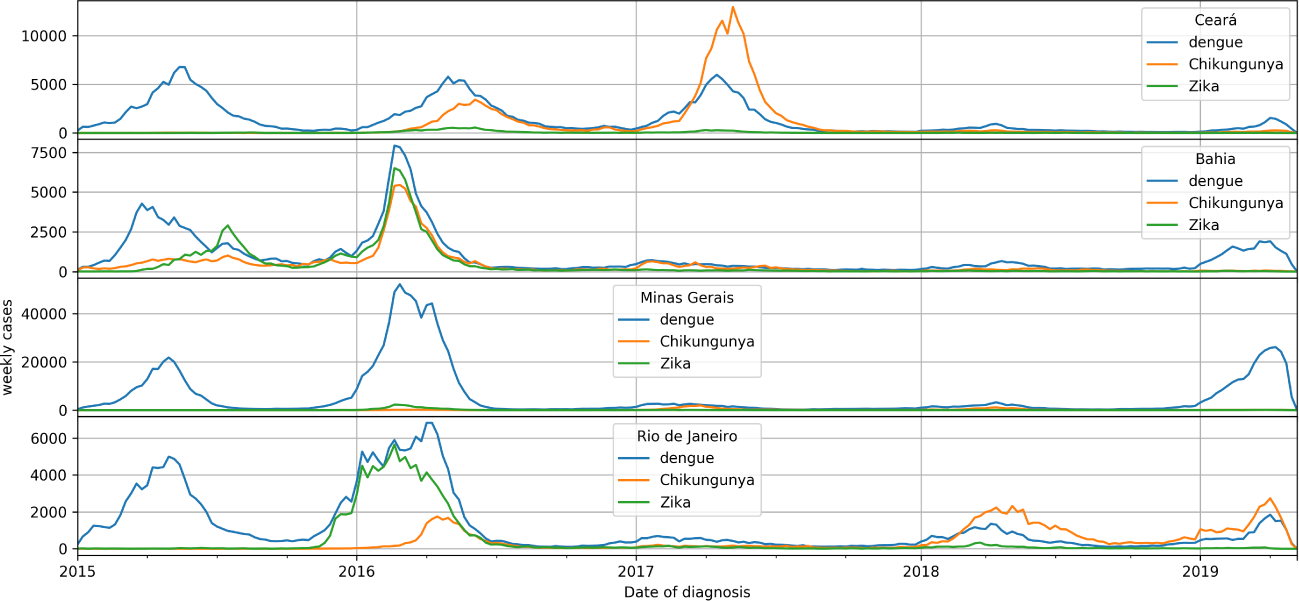
Weekly incidence for the state of Ceará, Brazil. Here we can see how the three arboviroses are correlated.

### Forecast models

Both LSTM and RQF models were trained to predict 4 weeks ahead of the last data point(*w*_*t*+4_). Both models use 4 weeks of historical data to generate forecasts. Forecasts are done in a rolling window fashion. Both models use as predictors, the following series: number of cases, Effective reproduction number (*R*_*t*_),Temperature, Humidity and Atmospheric pressure. Besides relying on local data, the forecast models also includes the same set of series from other cities belonging to a cluster of “epidemiologically similar” cities, defined by hierarchical agglomerative clustering based on the correlation distance between each city, i.e., the higher the correlation between their incidence time series the closer they are [2].

#### LSTM

A LSTM model is a recurrent deep neural network model developed to handle predictions of time series [3]. We used a LSTM model with 3 LSTM units followed by 3 dropout layers. The model was trained for 300 epochs with early stopping using a mean-log squared-error (MLSE) loss function and a Nesterov Adam optimizer [4]. Besides playing an important role in preventing overfitting during training [5], the dropout layers were also used in the prediction step to generate an ensemble of LSTM networks. This allows us to estimate confidence intervals by computing the relevant statistics from several predictions sampled from the ensemble [6]. Figure 2 shows dengue forecasts by the LSTM model for the city of Fortaleza. The black and red lines represent the incidence and the predictions of neural network respectively. The shaded blue area gives the corresponding 95% confidence interval for the predictions estimated from dropout sampling. The predictions curve (i.e, the red line in the figure) is simply the median of predictions computed with dropout sampling. The dashed green vertical line in the figure marks the boundary between training and test data. As can be seen from the figure, the model performs well even a few years in the future.

**Fig 2.**
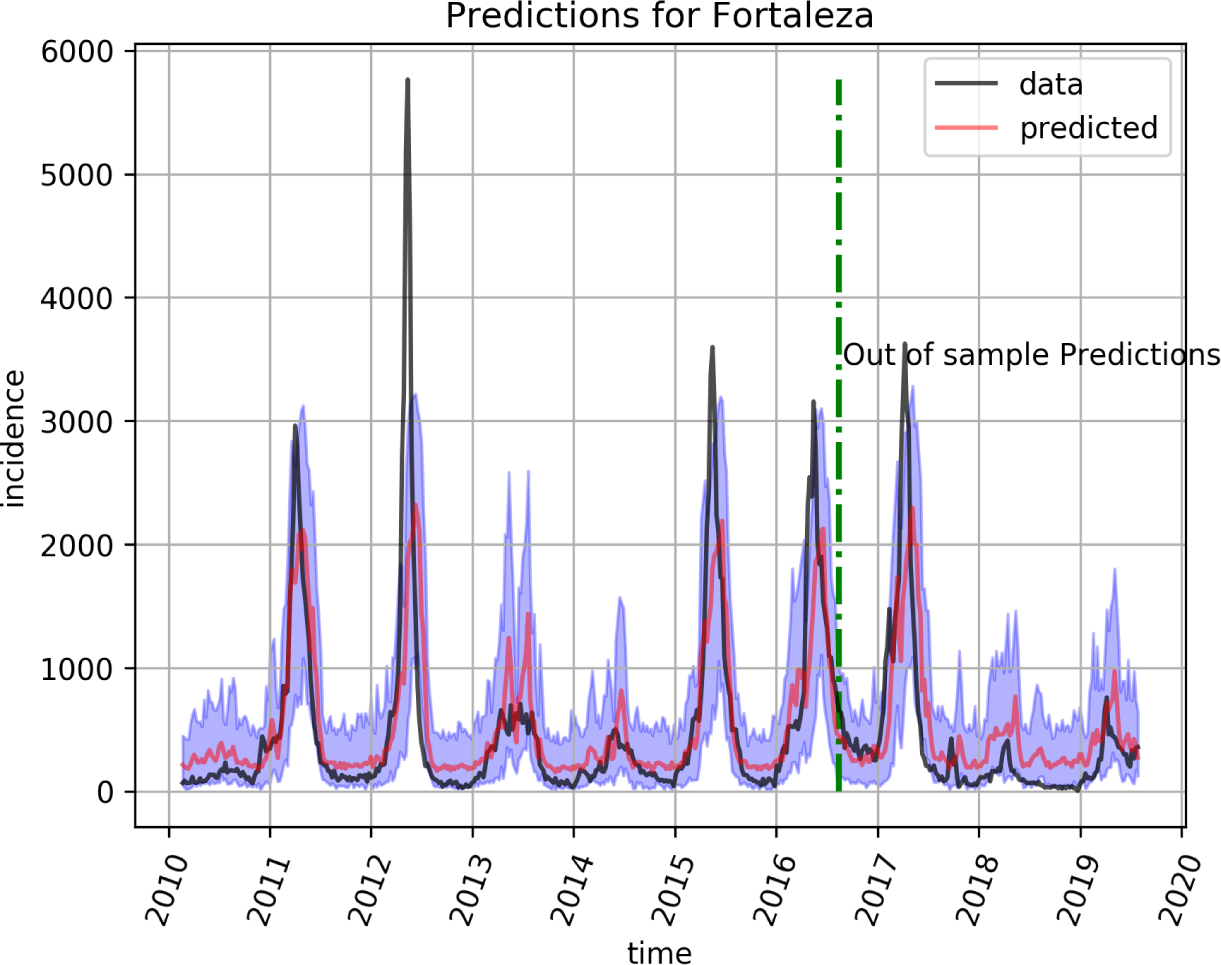
LSTM forecast for dengue incidence (black line) Prediction (red line) with 95% interval for the city of Fortaleza. Every point in the red line corresponds to a prediction from 4 weeks before.

#### Random Quantile Forest (RQF)

Random Forest models calculate an ensemble of regression trees from random subsets of data. RQF models are an extension to regular random forests in which the full conditional distribution of *Y* given *X* = *x* is calculated. As a result, it is a non-parametric, consistent and accurate way to determine conditional quantiles from high-dimensional predictors [7]. Let *A* = [*a*_*ij*_] be an array containing the *𝒟* = 4 most recent observations from each predictor series. Thus the regression model can be simply represented by

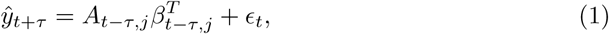

where *τ ∈* (1 … 4) and *j ∈* (1..*n*) where n is the number of predictors.

The full model includes the 5 predictor series for the city being predicted plus the same set from every other city belonging to its cluster.

Figure 3 shows dengue forecasts for Rio de Janeiro by the RQF model.

**Fig 3.**
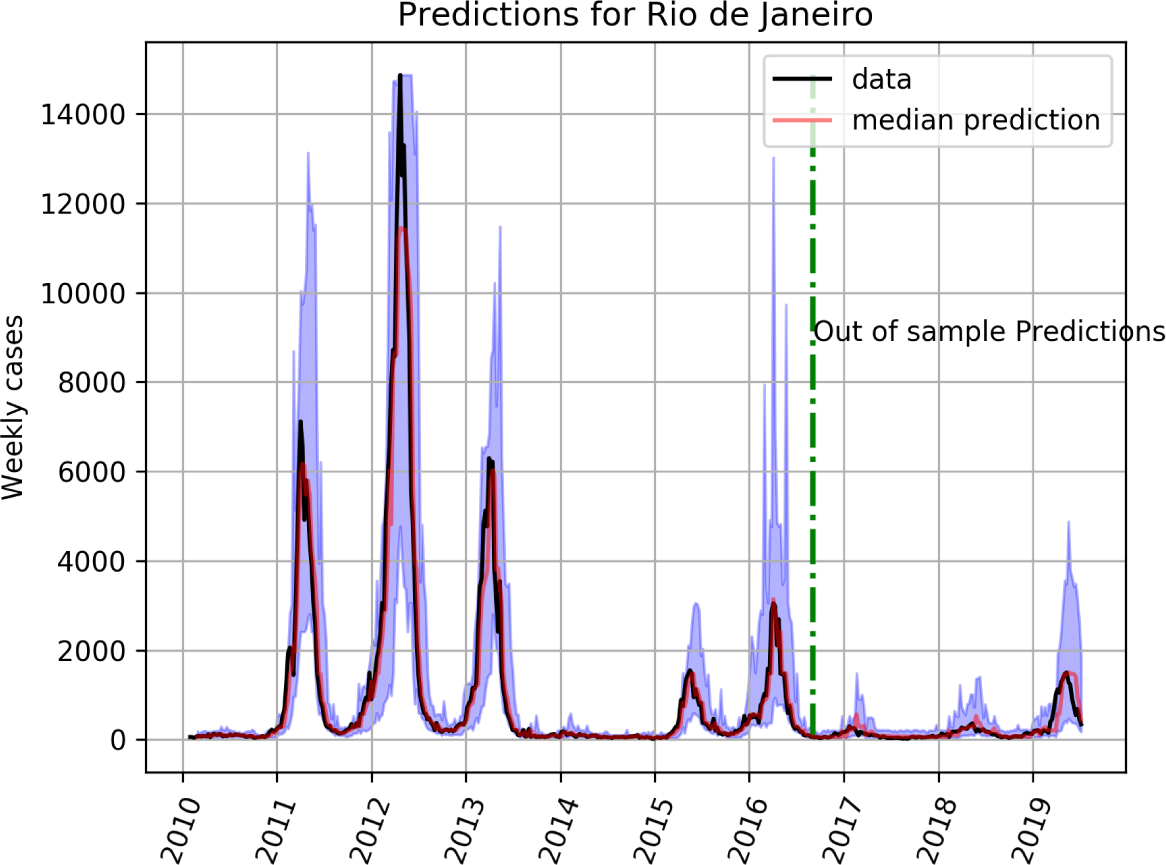
Baseline forecast for dengue in Rio de Janeiro. Random quantile forest model trained on data from 2010 to mid 2016. Red line is the median prediction, with 95% intervals in light purple.

### Model fitting and usage

Both models where trained with dengue and environmental data from 2010 to 2019. They were then saved and used for rolling predictions using Zika and Chikungunya series substituting for the dengue data. Predictions for Zika and Chikungunya, based on the models learned from the dengue data, were generated only from 2016 onward since before this year the two viruses were virtually absent from Brazil.

### Comparing models on a quantile scale

In order to allow the pooling of the prediction errors of different weeks *w* and cities *c* into a common scale, we transformed all the observations and predictions to their quantile value on the empirical cumulative distribution of historical incidences at each epidemiological week and city. Thus all errors become a difference between quantiles:

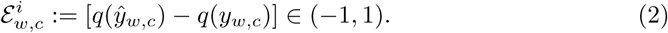

This quantile difference will give us part of the diagnostic. It will tell us how accurately the model predicted the actual week it is trained at. However, one may also be interested on how well the model has predicted the historical median. We will denote this second deviation by ξ := [*q*(*ŷ*_*w,c*_)*−*0.5] ∈ (*−*0.5, 0.5).

For the purpose of assessing the viability of applying transfer learning from dengue to Chikungunya and Zika we will consider only the first error, *ε*.

## Results

Figures 2 and 3 show rolling predictions 4 weeks ahead both in-sample and out-of-sample along with 95% predictive intervals, for dengue in Fortaleza and Rio de Janeiro, respectively. They illustrate the predictive accuracy of both models. These serve as baseline for the performance of cross-forecasting.

The performance of RQF for Chikungunya is shown in figure 4 for Rio de Janeiro and on figure 5 for Fortaleza. For Zika the predictions of the RQF model for the city of Fortaleza can be seen on figure 9.

**Fig 4.**
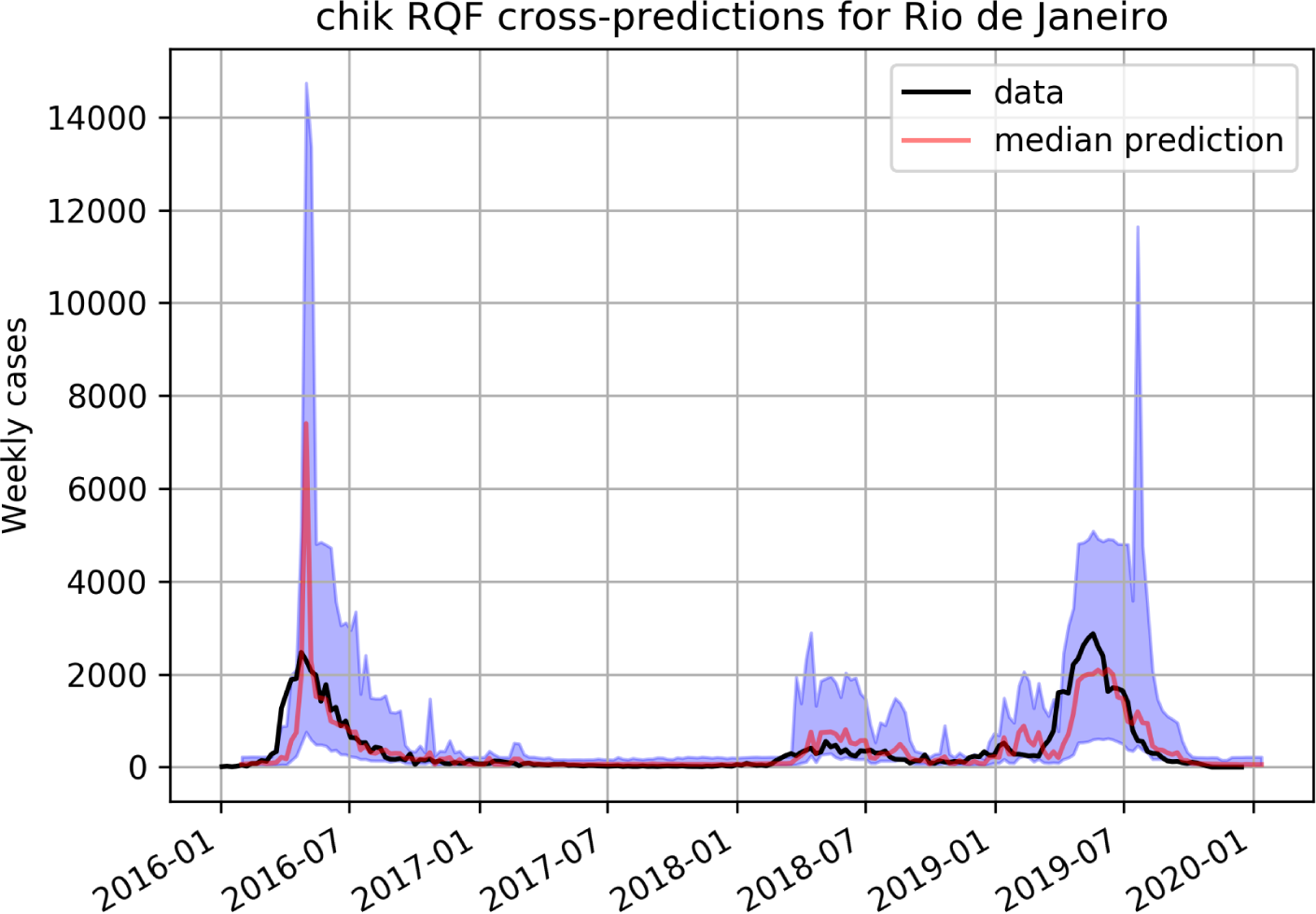
Predicting Chikungunya using a RQF model trained on dengue data in Rio de Janeiro.

**Fig 5.**
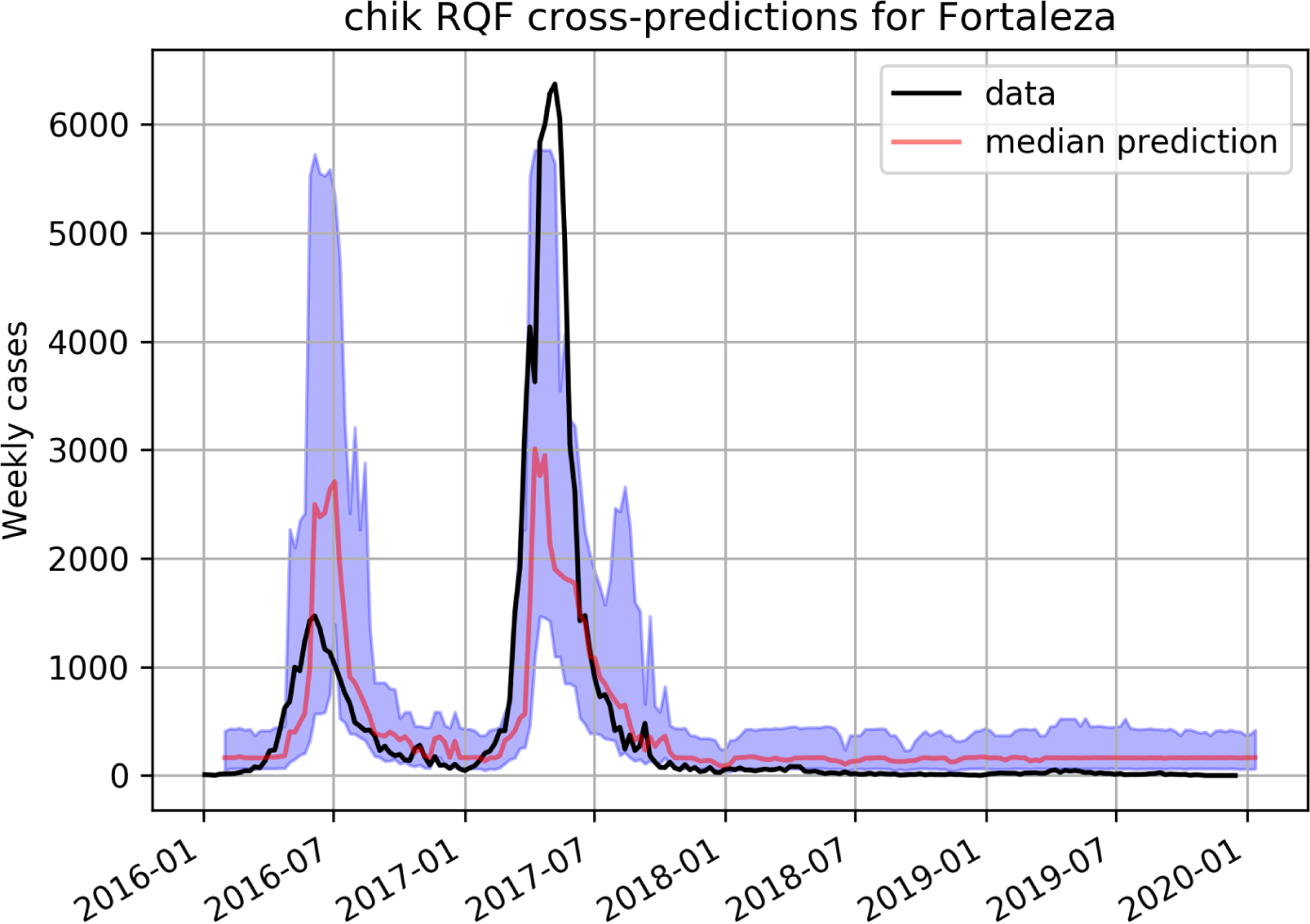
Predicting Chikungunya using a RQF model trained on dengue data in Fortaleza.

LSTM cross-prediction results for Chikungunya are shown in figures 6 and 7 for Rio and Fortaleza respectively, and on figure 8 for Zika.

**Fig 6.**
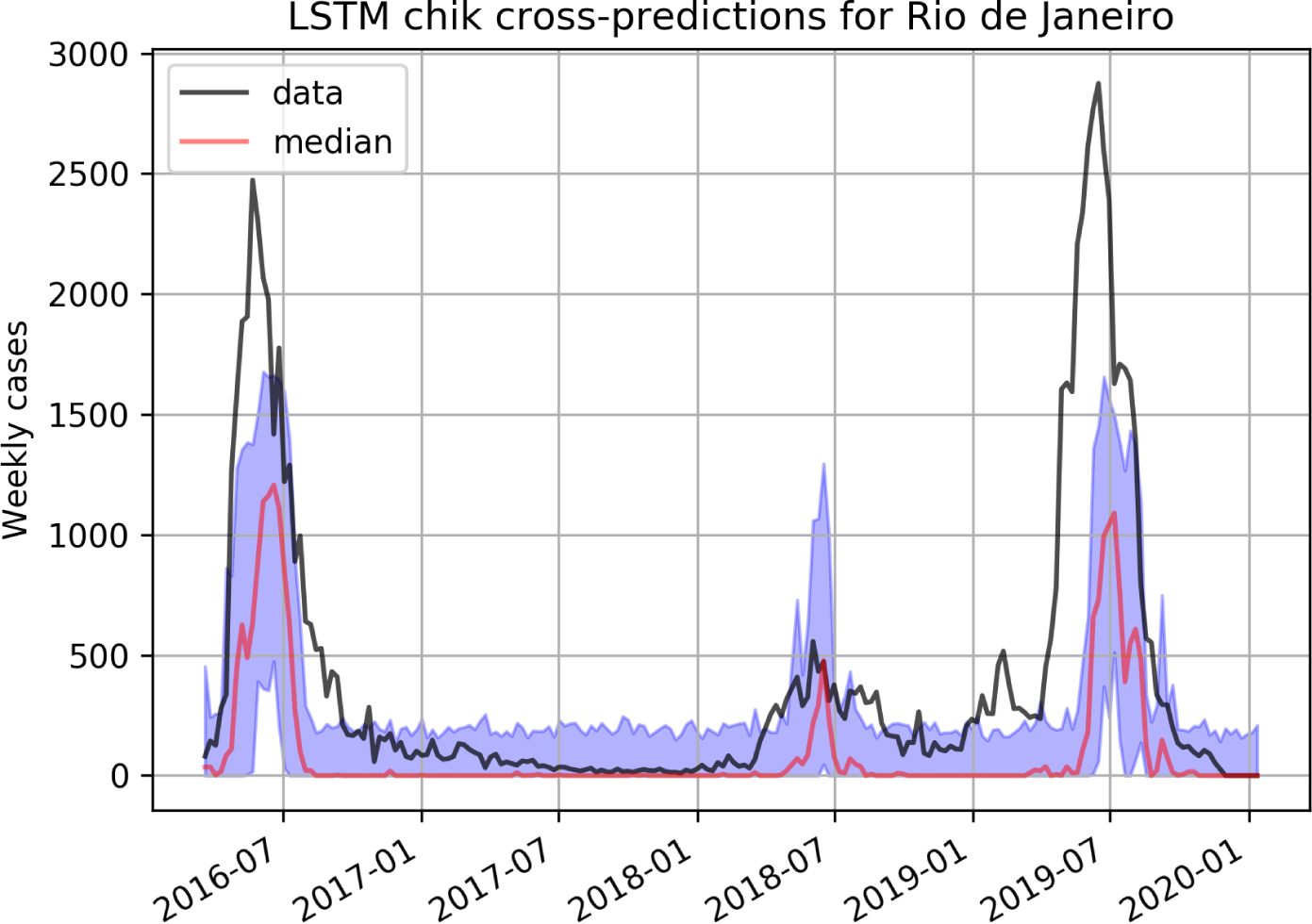
Predicting Chikungunya using a LSTM model trained on dengue data in Rio de Janeiro.

**Fig 7.**
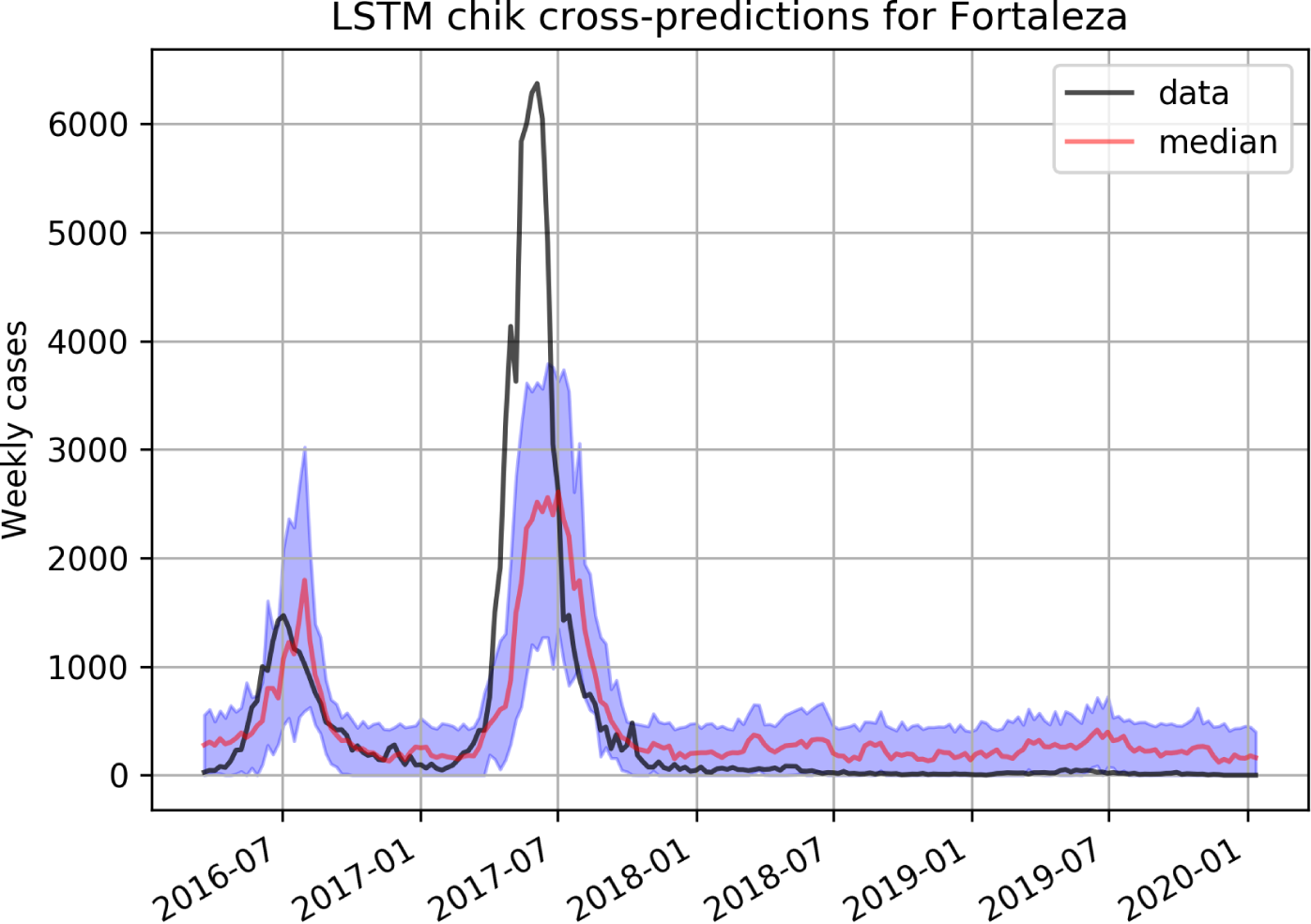
Predicting Chikungunya using a LSTM model trained on dengue data in Fortaleza.

**Fig 8.**
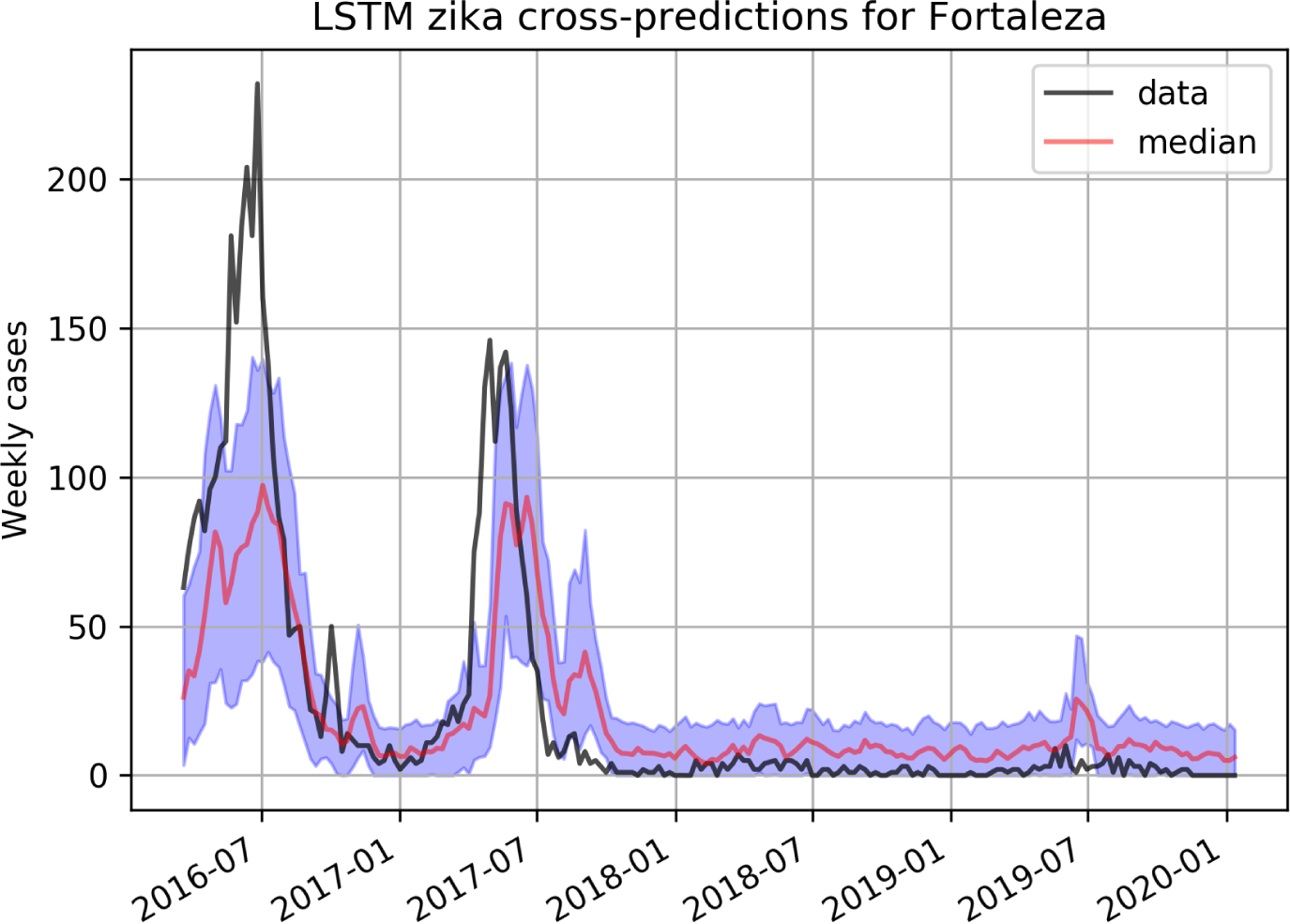
Predicting Zika using a LSTM model trained on dengue data in Fortaleza.

**Fig 9.**
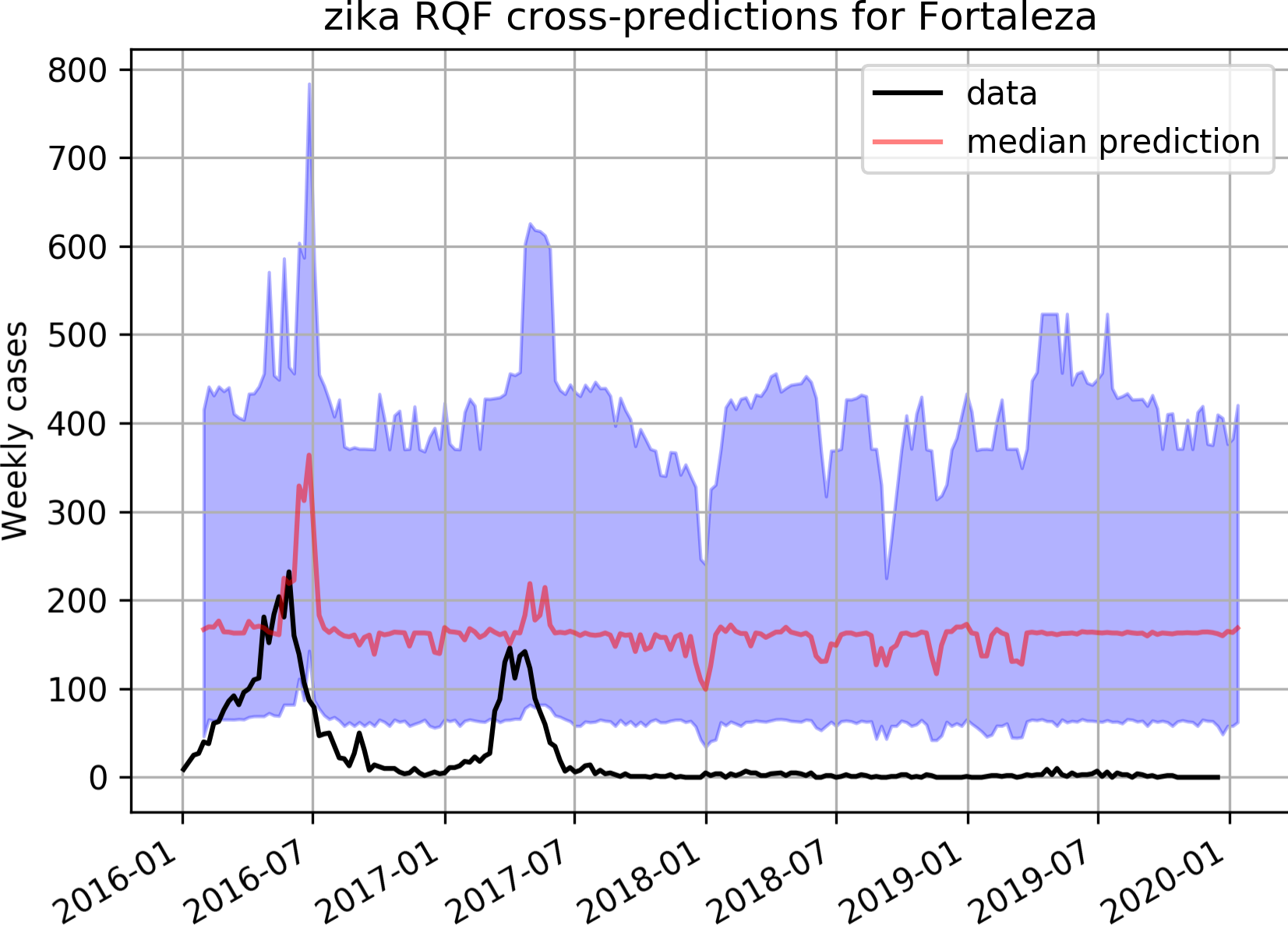
Predicting Zika using a RQF model trained on dengue in Fortaleza

Table 1 summarizes the cross-predictive errors for both models on both Zika and Chikungunya, for each city.

**Table 1.** **Distribution of predictive errors(*ε*, from equation 2)** for the LSTM and RQF models. The table shows the 1st quartile, **median**, and 3rd quartile, respectively, for each model, city and disease.

| Target | RQF |  |  | LSTM |  |  |
| --- | --- | --- | --- | --- | --- | --- |
|  | Fortaleza |  |  |  |  |  |
| Chikungunya | -0.38 | -0.24 | -0.04 | -0.45 | -0.29 | -0.02 |
| Zika | -0.59 | -0.47 | -0.35 | -0.36 | -0.24 | -0.05 |
|  | Rio de Janeiro |  |  |  |  |  |
| Chikungunya | -0.12 | -0.02 | 0.04 | 0.22 | 0.35 | 0.48 |
| Zika | -0.59 | -0.47 | -0.35 | N/A | N/A | N/A |

Another representation of the cross-prediction errors can be found on figures 10, 11, 12 and 13 for Chikungunya on both Rio de Janeiro and Fortaleza. These figures plot quantiles of the prediction values, *q*(*ŷ*_*w,c*_) against the quantiles of the observations, *q*(ŷ_*w,c*_). All weeks are aggregated on a single plot, so when we observe that a given model disagrees with the observations on a given percentile, we are not referring to a particular week, or magnitude. In other words, if the predicted quantile is higher than the observed quantile, when both are below the median, the prediction is actually a better approximation to the historical median than the observed value whenever it is below the historical expectation.

**Fig 10.**
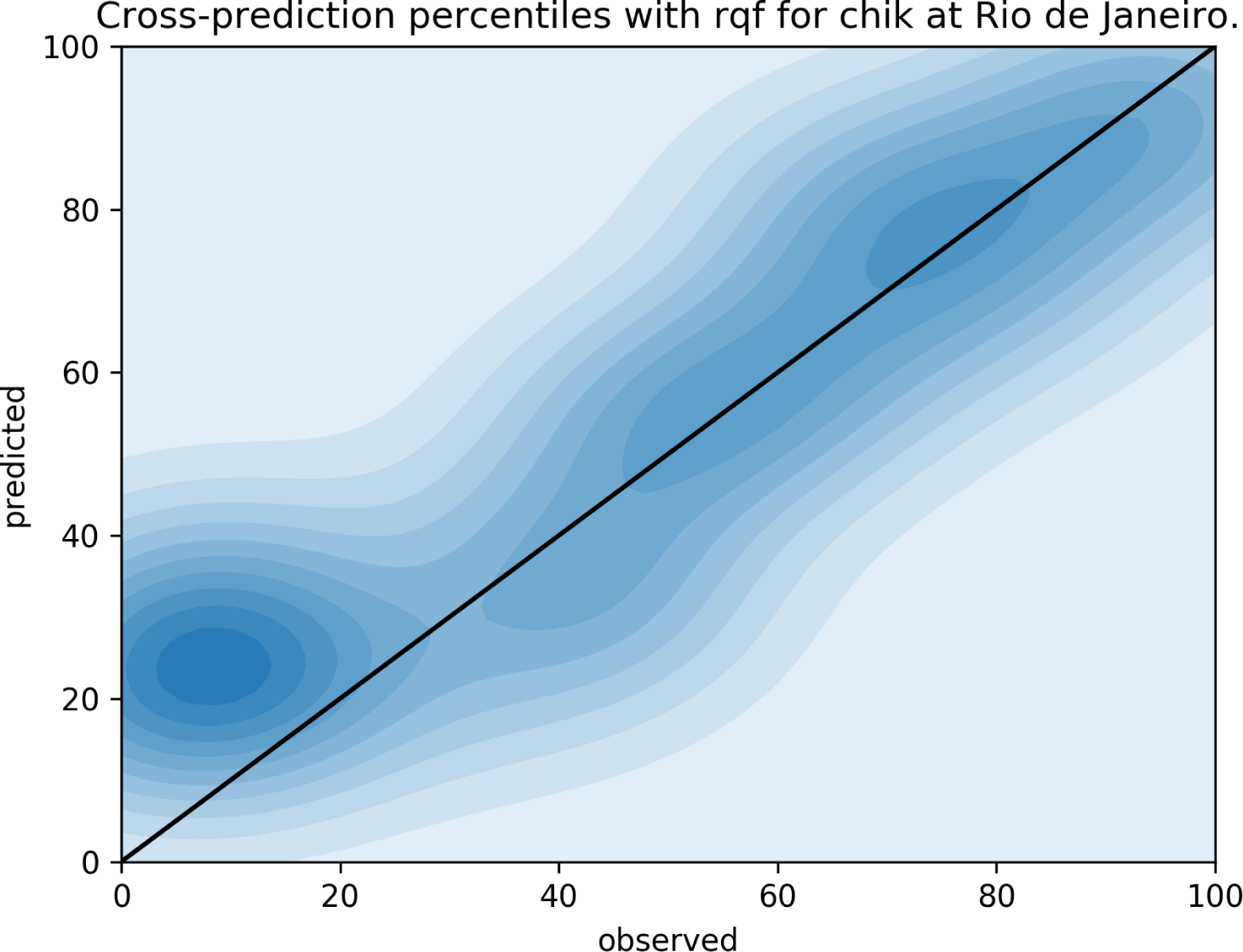
QQplot for observed (x-axis) and predicted (y-axis) weekly incidences in the city of Rio de Janeiro, cross-predicting Chikungunya from the dengue RQF model. Here we can see that when the observed incidence is below the 20th percentile, the RQF model tends to predict closer to the historical median overshooting the observed value for the week.

**Fig 11.**
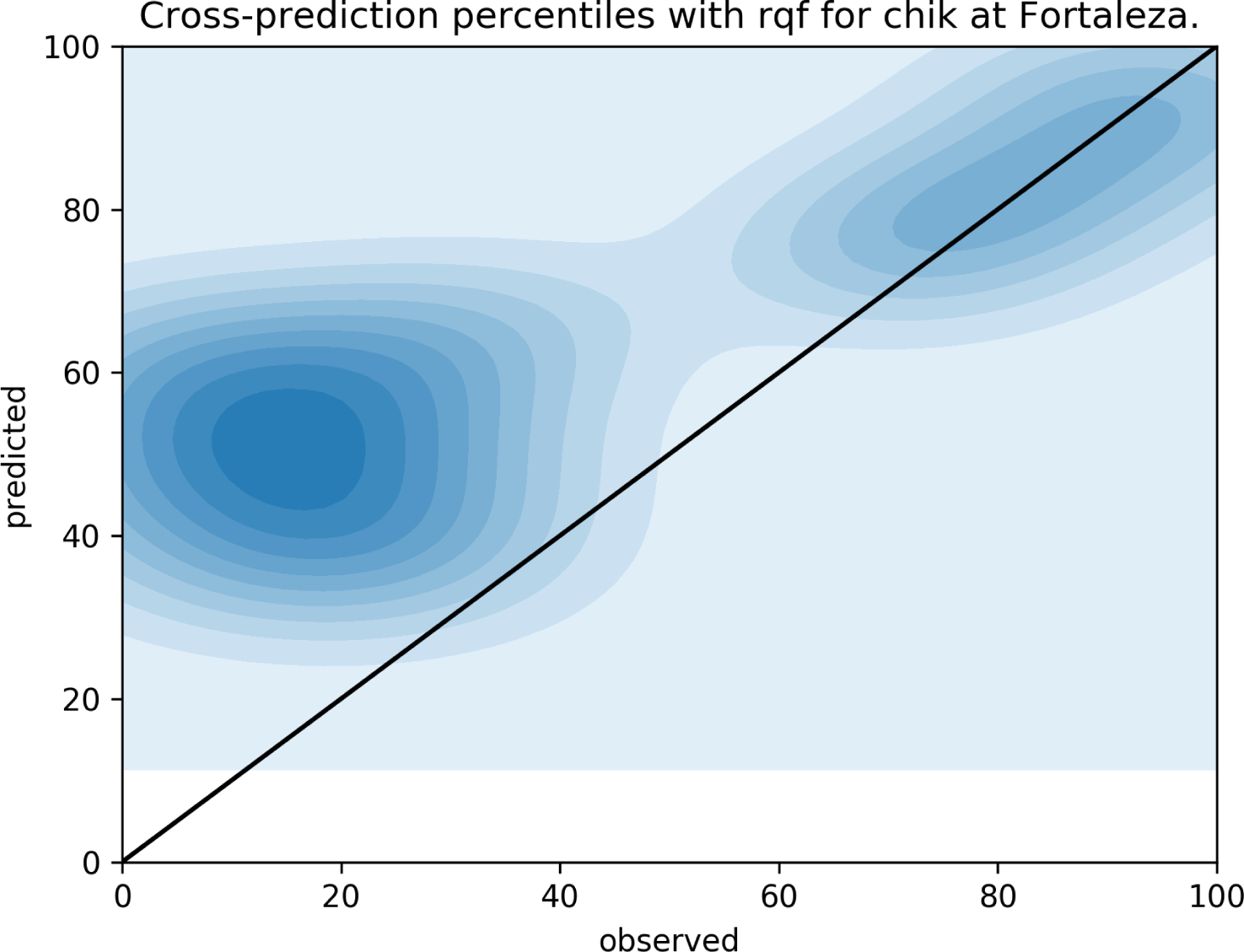
QQplot for observed (x-axis) and predicted (y-axis) weekly incidences in the city of Fortaleza, cross-predicting Chikungunya from the dengue RQF model. Here we note that the model’s predictions are right on the historical median whenever the observations are below this expected level. Whenever the observed incidence is higher than the expected incidence, the model correctly predicts these values (top-right of the plot).

**Fig 12.**
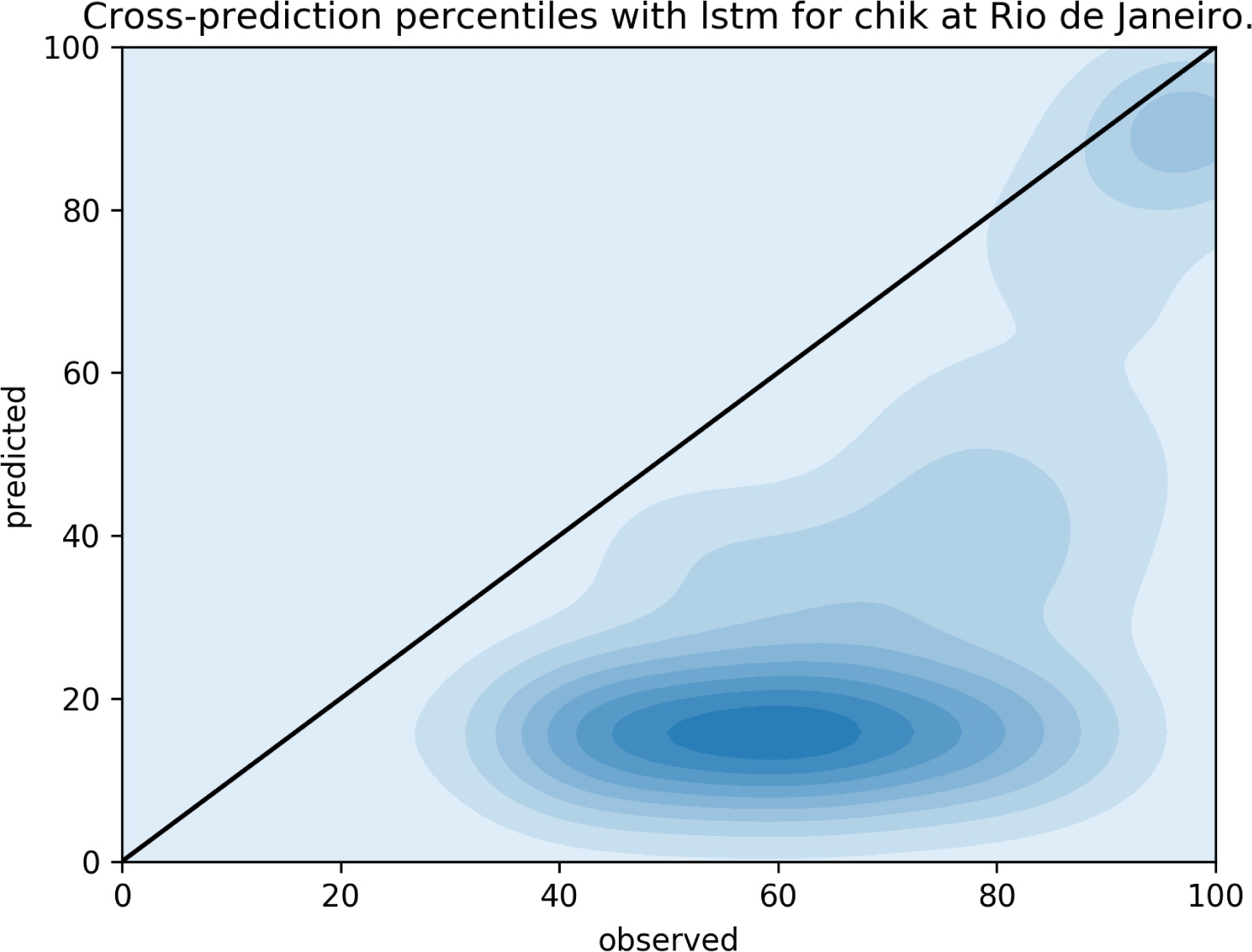
QQplot for observed (x-axis) and predicted (y-axis) weekly incidences in the city of Rio de Janeiro, cross-predicting Chikungunya from the dengue LSTM model. In this case, the LSTM model seems to consistently underestimate the observed incidence.

**Fig 13.**
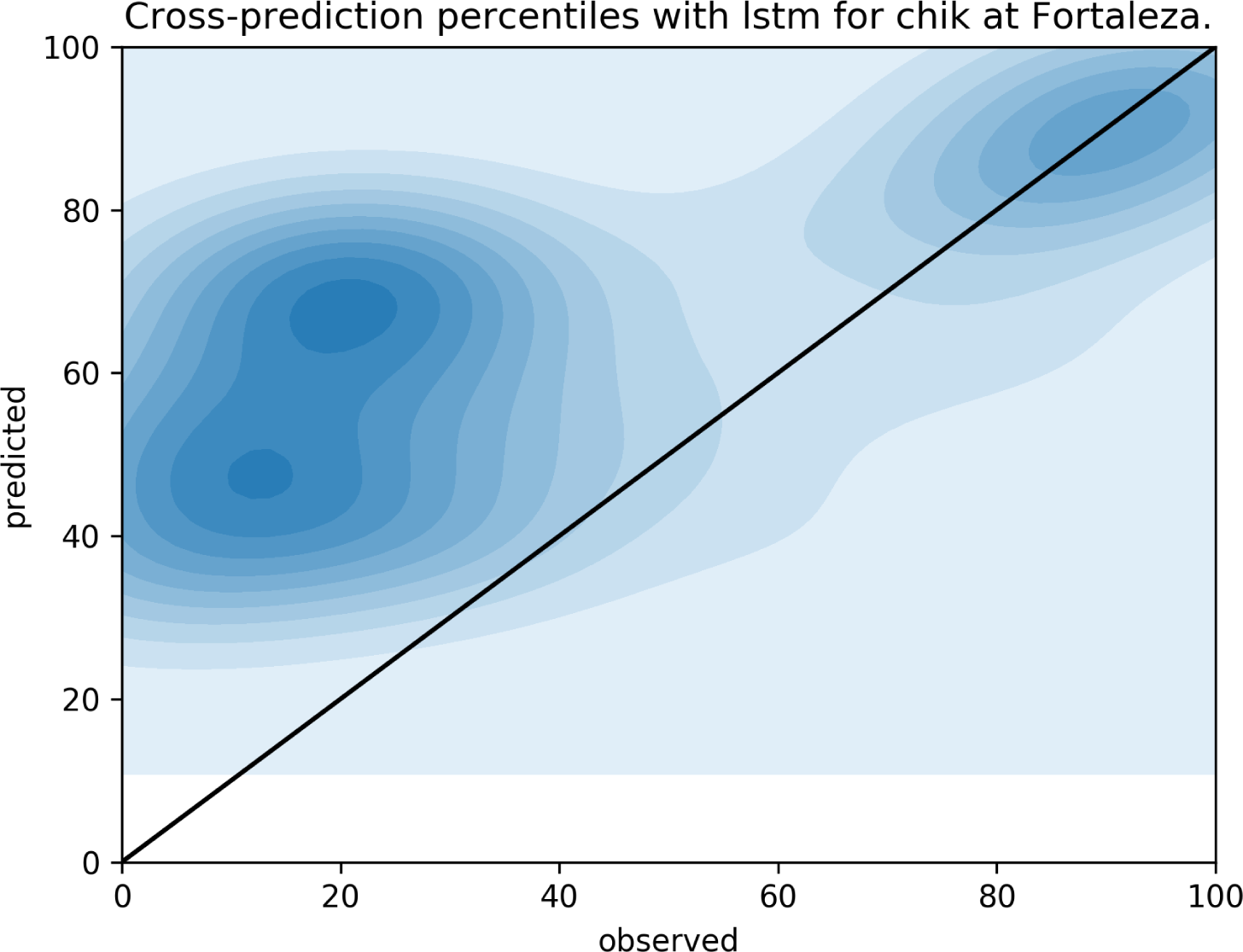
QQplot for observed (x-axis) and predicted (y-axis) weekly incidences in the city of Fortaleza, cross-predicting Chikungunya from the dengue LSTM model. In the city of Fortaleza the LSTM model seems to go for the historic median when the observed values are below the percentile 20 of the historical distribution. But the model and observations tend to be more in agreement when observations are at or above the historical median.

## Discussion

As shown in the results section, transfer leaning between dengue incidence forecast models and other arboviroses transmitted by the same vector can be an interesting approach. Typically, in transfer learning applications, the previously trained models get retrained with the new data set, to help the model combine the specifics of the new data set with the previously learned problem. However, for this application, we skipped this retraining step in order to retain good-sized data sets with which to test the models. Had we used the shorter Zika and Chikungunya time series to retrain the models, we would limit ourselves to provide only in-sample forecasts. On the other hand, by making this decision we imposed a much harder challenge to the models, namely predicting Zika and Chikungunya, having only been trained on dengue time series.

For this paper we have chosen two brazilian capitals, Fortaleza, in the northeast of the country and Rio de Janeiro, in the southeast. These cities are separated by approximately 2500 kilometers, and are under quite different climate regimes. This choice reflects our desire to test the models in places where the diseases are under very different environmental influences. This is apparent on the results, where we see that the models don’t perform equally well when cross-predicting in each city.

Furthermore, we should consider what the differences in performance mean for each individual city when using a single model to predict weekly incidences of different arboviroses. Indeed, when using a model trained on the dengue data set of a fixed city to predict Zika or Chikungunya weekly incidences for the same city, the environmental predictor variables are kept the same for all three diseases and therefore the differences in the predictions reflect the unique epidemiological dynamics of each arbovirosis at that observed moment. Therefore, the method we presented here can also shed light on how different the epidemiological dynamics of Zika and Chikungunya are as compared to that of dengue. This is particularly relevant since it may indicate that other modes of transmission may be present for arboviroses that behave very differently from dengue, as is already known to be the case of Zika, which has confirmed cases of sexual transmission [8].

## Conclusion

Cross-disease forecasting models like the ones presented here are important because they allow us to take advantage from longer historic records from other diseases which share similar transmission mechanisms and environmental determinants.

The results presented here expectedly show more uncertainty associated to cross-forecasts when compared to forecasting the disease they were trained on.

Nevertheless the models show reasonable accuracy when cross-predicting Chikungunya (figures 4, 5, 6 and 7), perhaps more than the same models would be capable of if they were trained on the scarce available data for Chikungunya.

For Zika, cross-forecasts based of the RQF model did not work as well (figure 9). Perhaps the fact that Zika can also be transmitted sexually [8] makes its epidemiological dynamics sufficiently different from dengue to make cross-predicting ineffective. The LSTM model showed similar performances when compared to RQF for Chikungunya (figure 7). But for Zika (figure 8) the LSTM model performed much better than RQF for cross-predictions.

Machine learning models have shown great potential for infectious disease forecasting. However, long enough time series, essential to train such models, are not easy to come by. In this work we have shown the potential of cross-disease forecasts, for diseases with similar transmission mechanisms. Despite the encouraging initial results there is still plenty of room for improvements to the development and application of such models.

## Data Availability

Data is available through the Infodengue API

## Notes

### Competing Interest Statement

The authors have declared no competing interest.

### Funding Statement

No Funding was received for the realization of this work.

## References

1. Codeco C, Coelho F, Cruz O, Oliveira S, Castro T, Bastos L. Infodengue: A nowcasting system for the surveillance of arboviruses in Brazil. Revue d’Épidémiologie et de Santé Publique. 2018;66:S386.

2. Mussumeci E. A machine learning approach to dengue forecasting: comparing LSTM, Random Forest and Lasso. School of Applied Mathematics, Fundação Getulio Vargas; 2018.

3. Hochreiter S, Schmidhuber J. Long short-term memory. Neural computation. 1997;9(8):1735–1780.

4. Dozat T. Incorporating nesterov momentum into adam; 2016.

5. Srivastava N, Hinton G, Krizhevsky A, Sutskever I, Salakhutdinov R. Dropout: a simple way to prevent neural networks from overfitting. The journal of machine learning research. 2014;15(1):1929–1958.

6. Gal Y, Ghahramani Z. Dropout as a bayesian approximation: Representing model uncertainty in deep learning. In: international conference on machine learning; 2016. p. 1050–1059.

7. Meinshausen N. Quantile regression forests. Journal of Machine Learning Research. 2006;7(Jun):983–999.

8. Coelho FC, Durovni B, Saraceni V, Lemos C, Codeco CT, Camargo S, et al. Higher incidence of Zika in adult women than adult men in Rio de Janeiro suggests a significant contribution of sexual transmission from men to women. International Journal of Infectious Diseases. 2016;51:128–132.

